# Traditional Chinese Medicine Research Activity in Switzerland. A Systematic Review and Bibliometric Analysis

**DOI:** 10.1101/2024.07.10.24310251

**Authors:** Bingjun Chen, Xiaoying Lyu, Koryna Lottenbach, Chantal Zeier, Dominic Jaminet, Sarangoo Solongo, Hummel Timo, Wicker Talia, Ahmet Sevik, Xiaying Wang, Michael Hammes, Yiming Li, Saroj K. Pradhan, Michael Furian

## Abstract

**Importance:** Traditional Chinese Medicine (TCM) becomes popular in Switzerland, however, Swiss TCM research activity and scientific output have not been investigated.

**Objective:** To describe the Swiss TCM research activities and main health conditions studied.

**Data Source:** A systematic literature search of PubMed and Embase was performed from database inception to December 31^st^, 2023.

**Study Selection:** Articles describing a TCM-related therapy modality in humans with at least one author affiliated with a Swiss institution.

**Data Extraction:** Primary and secondary outcomes, as well as study and author characteristics were extracted from included articles.

**Main Outcomes and Measures:** The main outcome was the main health condition studied. Secondary outcomes are the total number of articles published over time, the TCM therapies used, and the Swiss institutions involved.

**Results:** Of the 223 articles included, 68.2% of published articles originated from the top 3 of 73 (4.1%) Swiss institutions, namely the University of Zurich (32.3%), University of Bern (30.0%) and University of Basel (7.6%). Overall, 116 (52.0%) articles described original studies including 36 (31.0%) articles reporting findings from randomized clinical trials, 29 (25.0%) from cross-sectional studies, 20 (17.2%) from prospective cohort studies, 19 (16.4%) from case reports and 12 (10.3%) from retrospective studies. The top health categories studied were Pain Management (16.4%), Psychology and Behaviour (8.6%), Neurology (6.9%), and Oncology (6.9%). The most used TCM therapies were acupuncture or moxibustion (61.2%), combination of several treatments (15.5%), herbal medicine (10.3%), and Qi Gong or martial arts (9.5%).

**Conclusion and Relevance:** Until 2023, the total number of scientific TCM output by Swiss authors is steadily increasing but remains small. More effort to conducted TCM research and to elucidate the TCM therapy effects in Switzerland is warranted.

**Registration:** PROSPERO No. CRD42023432693.

## 1. Introduction

Complementary medicine (CM) is widely used in Switzerland. In a 2009 referendum, two-thirds of Swiss voters approved a new article in the Federal Constitution that recognises CM by the authorities. Since 2009, Switzerland has seen a steady increase in the use of CM, and according to data from the Swiss Health Survey of 2007 and 2012, 23.0% and 25.0%, respectively, of the population aged 15 and older had used at least one CM method in the preceding 12 months.[1] Another survey by the Swiss Federal Statistical Office in 2017 confirmed the steady increase of CM and reported a prevalence of CM use of 28.9%.[2] In 2024, four CM therapies (anthroposophy, homeopathy, herbal medicine, and traditional Chinese medicine [TCM]) are fully covered by mandatory basic health insurance if delivered by a certified physician or by complementary insurance when delivered by certified therapists.[3]

TCM has gained increasing popularity and includes modalities such as acupuncture, Chinese herbal medicine, Tuina massage, dietary therapy, and physical activities such as Tai Qi and Qi Gong. Randomized clinical trials, systematic literature reviews, and meta-analyses have provided evidence to support the efficacy of several TCM modalities in managing various diseases.[4–7] Moreover, TCM has the potential to provide relief for certain health conditions, which conventional medicine yet struggles to treat or yield medication side-effects.[8, 9]

However, it is important to note that most of the studies and evidence for TCM treatment effects originate from China and substantial differences in TCM education, access to Chinese herbs, use of TCM therapies or limited access to the Chinese literature might limit the transferability and generalizability of TCM therapy effects across the globe. Moreover, the strength of TCM lays in personalized medicine rather than standardized procedures, another factor which requires further attention when quantifying therapy effects and sustainability. To minimize these confounding factors, including cultural differences, findings from TCM studies in Switzerland and conducted by Swiss research institutions are highly warranted because their findings on the efficacy of various TCM therapies might be most applicable to the Swiss population and its related healthcare system.

Therefore, the aim of this systematic literature review was to summarize the Swiss TCM research activities, studied health conditions and chosen TCM therapies, to understand the current level of research activity, and to cost-effectively guide future TCM research directions in Switzerland.

## 2. Methods

This systematic literature review follows the Preferred Reporting Items for Systematic Reviews and Meta-Analyses (PRISMA) guidelines (**Supplemental Table S1**)[10] and is registered at PROSPERO [number: CRD42023432693].

### 2.1. Literature search

A systematic literature search was conducted on PubMed and Embase from database inception to December 31st, 2023. The applied search term was “*(TCM or Traditional Chinese Medicine or Chinese herbal medicine or Acupuncture or Electroacupuncture or Moxibustion or Tuina or Tai Qi or Taiji or Taichi or Qigong) AND Switzerland[affil]”.* Titles and abstracts of the records were screened, and the full texts were obtained if they met the inclusion criteria.

### 2.2 Data extraction

Data were extracted by 3 investigators (K.L, C.Z, D.J) and independently by the author B. C. using a standardized, pre-piloted form for all studies meeting eligibility criteria for inclusion. When uncertainties about eligibility arose, an agreement was found through discussions.

In all included studies, the following data were extracted:

#### Data related to the article

year of publication; PMID; type of article; research topic; keywords; study design; medical journal metrics;

#### Data related to the population

sample size; demographics including age, gender and primary disease/health condition; treatment modality;

#### Data related to the Swiss author and institution

number of total authors; Swiss author position (first, co-, last author); Swiss institution.

### 2.3 Study selection criteria

Articles describing research in humans and written in German, French, English, or Chinese with at least one author affiliated to a Swiss institution were selected for the final analysis.

### 2.4 Statistical analysis

Aggregated data from all included studies are descriptively presented in tables by numbers and proportions or means ± standard deviation. To analyse and characterize the primary health condition of the retrieved articles, health condition categories were defined. For analyses related to the Swiss institutions and authors, we unified different expressions of the same institution and merged different departments of the same institution under a single name, i.e. (1) Lausanne University Hospital and University of Lausanne; (2) University Hospital of Bern, University Women’s Hospital Bern, and University of Bern; (3) University Hospital Zurich, Children’s Hospital Zurich, and University of Zurich; (4) University Hospital of Geneva and University of Geneva; (5) University Hospital of Basel, University Children’s Hospital of Basel and University of Basel; (6) Lausanne University Hospital and University of Lausanne; (7) Swiss Paraplegic Centre, Swiss Paraplegic Foundation and Swiss Paraplegic Research; and (8) Hirslanden Zurich AG, Hirslanden Clinic Aarau, and Hirslanden Clinics. Subgroup analysis was performed to gain further insights into the content of original articles containing information from case reports, retrospective, cross-sectional and prospective studies as well as randomized clinical trials. To understand the frequency and quantity of collaborations between Swiss institutions, we performed social network analysis. Data analysis was conducted in R software (version 4.3.2) and VOSviewer software (version 1.6.20) was used to visualize the network relationship among Swiss research institutions.

## 3. Results

### 3.1 Study Selection

A total of 918 studies were identified through the literature search on PubMed and Embase. After assessing all studies, a total of 223 studies were included in this systematic literature review (**Figure 1, Supplemental Table S2**).

**Figure 1:**
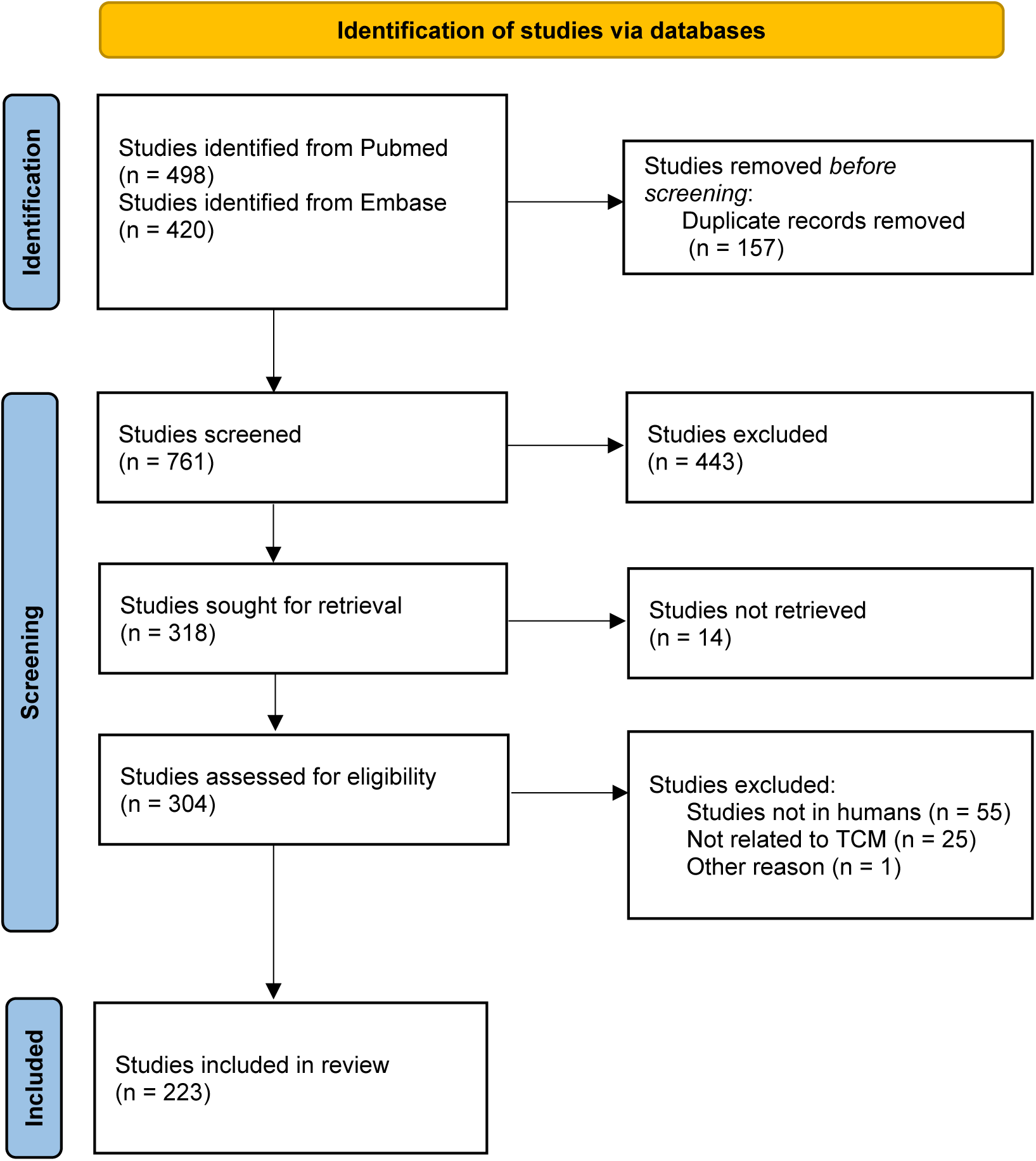
Study selection flow chart. TCM, Traditional Chinese Medicine.

### 3.2 Characteristics of included articles

Among the 223 included articles, 116 (52.0%) were categorized as *original articles*, 45 (20.1%) as *systematic reviews*, 39 (17.5%) as *narrative reviews*, 3 (1.3%) as *letters to the editor*, and 20 (9.0%) as *other type of articles* (**Table 1**). Overall, 148 of 223 (66.4%) articles were published in Science Citation Index Expanded (SCIE) journals with a mean ± SD journal impact factor percentile in the year of publication of 65.9 ± 22.2.

**Table 1.** Overview of the included articles.

|  |  |
| --- | --- |
| **Article information** |  |
| Total number of articles, n | 223 |
| Total number of articles in SCIE journals | 148 (66.4%) |
| JIF Percentile, %<sup>1</sup> | 65.9 ± 22.2 (range 4.2 to 99.8) |
| **Swiss author information** |  |
| Total number of Swiss researchers, N (% women) | 405 (37.8%) |
| Number of published articles by author | 1.5 ± 2.3 (range 1 to 39) |
| With First authorship, n (%) | 133 (59.6%) |
| With Co-authorship, n (%) | 154 (69.1%) |
| With Last authorship, n (%) | 146 (65.6%) |
| **Article type** |  |
| Original article, n (%) | 116 (52.0%) |
| Randomized clinical trial, n (%) | 36 (31.0%) |
| Cross-sectional study, n (%) | 29 (25.0%) |
| Prospective study, n (%) | 20 (17.2%) |
| Case report, n (%) | 19 (16.4%) |
| Retrospective study, n (%) | 12 (10.3%) |
| Systematic review, n (%) | 45 (20.1%) |
| Narrative review, n (%) | 39 (17.5%) |
| Others, n (%)<sup>2</sup> | 20 (9.0%) |
| Letter to the Editor, n (%) | 3 (1.3%) |

Values represent number and proportion or mean ± SD. <sup>1</sup>JIF, journal impact factor was defined as the JIF reported in the publication year of the article from the Journal Citation Reports from Clarivate. <sup>2</sup>Articles categorized as *others* included Guidelines, Essays, Study Protocols and Editorials. SCIE, science citation index expanded.

Between 1985 and 2000, only 8 articles met the inclusion criteria, while between 2001 and 2009, a total of 40 articles were published with Swiss author contributions (**Figure 2**). Between 2010 and 2023, in accordance with the TCM included in the health insurance reimbursement list in Switzerland in 2009, a substantial increase in the number of articles can be observed (78.5% of all articles). With 21 articles published in 2023, this year represents the climax in Swiss TCM research activity (**Figure 2**).

**Figure 2:**
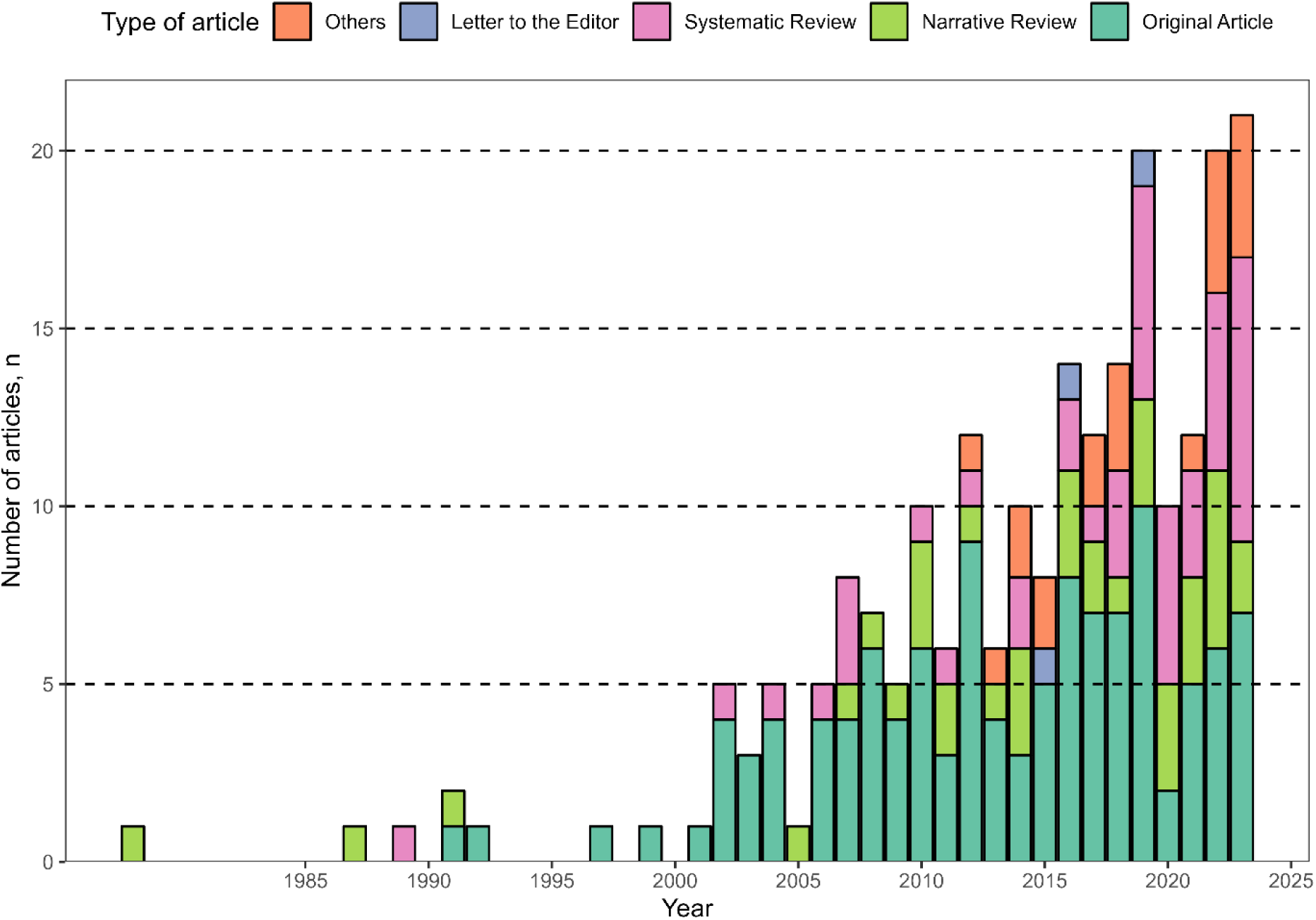
Number of published articles over time. This figure illustrates the absolute number of published articles between 1978 to 2023 and depicts the annual proportion of different article types. The category *Others* encompasses articles such as *Guidelines*, *Essays*, *Study Protocols* and *Editorials*.

Based on the 223 articles, 405 (37.8% women) individual authors affiliated with Swiss institutions were identified. Of these, 133 (59.6%) were first authors, 154 (69.1%) were co-authors, and 146 (65.5%) were last authors of the corresponding articles. On average, each Swiss author contributed to 1.5 ± 2.3 articles (**Table 1**).

### 3.3 Characteristics of original articles

One hundred and sixteen of 223 (52.0%) articles were categorized as original articles, including 36 (31.0%) articles originating from randomized clinical trials, 29 (25.0%) from cross-sectional studies, 20 (17.2%) from prospective cohort studies, 19 (16.4%) from case reports, and 12 (10.3%) from retrospective studies (**Table 1**).

**Table 2** provides information related to the original studies. The majority (81.9%) of studies were conducted in adults and comprised 17 different categories of the primary health condition. The most studied primary health conditions were categorized to *Pain Management* (16.4%), *Psychology and Behaviour* (8.6%), *Neurology* (6.9%) and *Oncology* (6.9%) (**Figure 3**).

**Figure 3:**
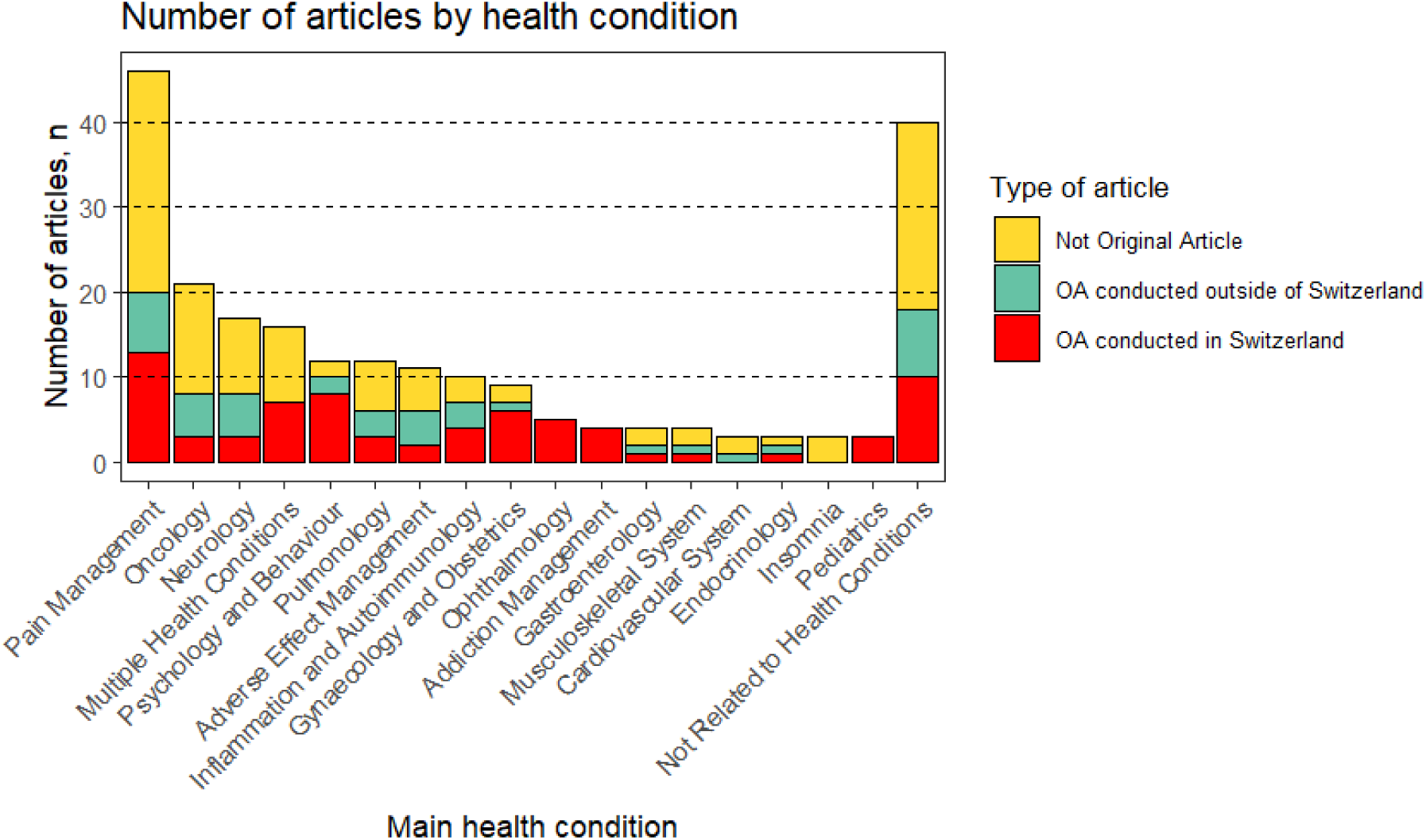
Articles stratified by primary health condition. This figure illustrates the number of articles categorized by the primary health condition described in the article. Articles categorized as *“Not related to health conditions”* focussed on clinical studies in healthcare professionals, on physiologic/mechanistic topics, on medical policies or other topics not related to a disease or symptom. “Original articles conducted in Switzerland” are defined as research projects carried out in the Swiss population. OA, original article.

**Table 2.** Characteristics of original articles.

| **Table 2. Characteristics of original articles** |  |
| --- | --- |
| Number of original articles, n | 116 |
| Number of study participants, median [IQR] | 60 [20 ; 212] |
| Number of women participants, median (%) | 33, 48.5% |
|  | [24 articles without information related to sex] |
| **Age category of study participants** |  |
| <18 years, n (%) | 4 (3.4%) |
| ≥18 years, n (%) | 95 (81.9%) |
| No age restriction, n (%) | 5 (4.3%) |
| Not reported, n (%) | 12 (10.3%) |
| **Applied TCM therapy** |  |
| Acupuncture with/without moxibustion | 71 (61.2%) |
| Combination of several treatments | 18 (15.5%) |
| Herbal medicine | 12 (10.3%) |
| Qi Gong or Martial Arts | 11 (9.5%) |
| Acupressure and Tuina | 2 (1.7%) |
| Modern use of acupuncture points<sup>1</sup> | 2 (1.7%) |
| **Country, in which the study was conducted** |  |
| Switzerland | 74 (63.8%) |
| Germany | 18 (15.5%) |
| China | 9 (7.8%) |
| International | 4 (3.4%) |
| Greece | 2 (1.7%) |
| France | 2 (1.7%) |
| Italy | 1 (0.9%) |
| Netherlands | 1 (0.9%) |
| Bulgaria | 1 (0.9%) |
| Austria | 1 (0.9%) |
| Not reported | 3 (2.6%) |

Values represent number and proportion or median and interquartile range. IQR, interquartile range; TCM, Traditional Chinese Medicine. <sup>1</sup>Two articles used electrical stimulation of the acupuncture point and laser acupuncture.

In the majority of articles, *acupuncture with/without moxibustion* was used, followed by a *combination of several TCM therapies* and *Qi Gong or Martial Arts* (**Table 2**).

### 3.4 Research activities by institution

According to the 223 articles, a total of 73 Swiss institutions active in TCM research were identified. The top five institutions with the largest number of articles are the University of Zurich with 72 (32.3%), the University of Bern with 67 (30.0%), the University of Basel with 17 (7.6%), the University of Geneva with 16 (7.2%), and the University of Lausanne with 13 (5.8%) (**Table 3**). Only 7 of 73 (9.6%) Swiss institutions published 7 or more articles, but collectively account for 79.8% of all eligible articles. **Figure 4** shows the relationship of the published literature network of each Swiss institution.

**Figure 4:**
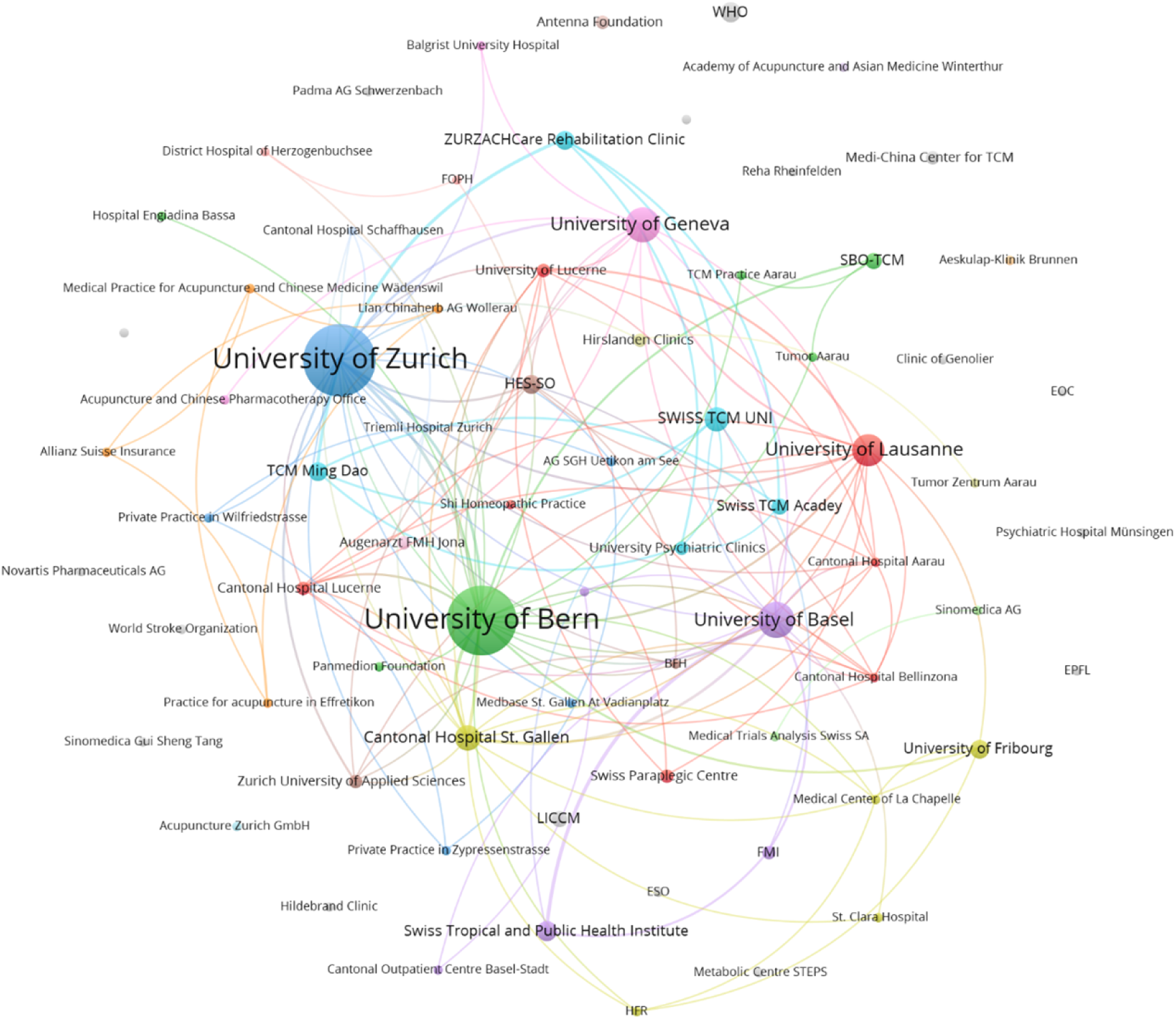
Network analysis of Swiss institutions active in TCM research. This figure illustrates a network analysis depicting the quantity of published articles by each Swiss institution and their collaborative relationships within articles. The circles and labels represent the number of published articles. The colour of the dots and connections indicate different clusters of collaboration networks.

**Table 3.** Swiss institutions active in TCM research.

|  |  |
| --- | --- |
| Number of Swiss institutions, n | 73 |
| **Top Swiss institutions, number of affiliations in included articles\*** |  |
| University of Zurich | 72 (32.3%) |
| University of Bern | 67 (30.0%) |
| University of Basel | 17 (7.6%) |
| University of Geneva | 16 (7.2%) |
| University of Lausanne | 13 (5.8%) |
| Cantonal Hospital St. Gallen | 8 (3.6%) |
| Swiss University of Traditional Chinese Medicine | 7 (3.1%) |

TCM, Traditional Chinese Medicine. \*several institutions can be affiliated within the same article.

## 4. Discussion

This registered systematic literature review analysed the TCM research activity in Switzerland from 1985 until 2023. Findings revealed a steady increase of TCM research activity after the decision to reimburse TCM through the Swiss health insurance system in 2009. Most research has been done by a few Swiss institutions focusing mostly on acupuncture and pain management, whereas other TCM therapies and health conditions were less prioritized. Although the observed positive TCM research activity trend, the absolute number of published articles (N = 223 since 1978) and original studies generating new knowledge (N = 116) remains low and might not match the broad use of TCM in Switzerland. Moreover, the number of original studies stagnates since several years and ≥10 published articles were only achieved in the fields of *Pain Management* and *Psychology and Behaviour*. These findings indicate that future medical research should prioritize TCM and its various subspecialties to better elucidate its optimal and complementary use in Western Medicine health systems.

The obtained findings and the prioritization of certain health conditions (**Figure 3**) are difficult to compare to the actual, real-world TCM use in the Swiss population and in TCM practices. Indeed, no quantitative data is available about the patient population and the distribution of diseases/symptoms presented and treated in TCM practices. This discrepancy requires further attention since a cross-sectional survey of 18,832 Swiss residents in 2017 showed that 7.0% used herbal medicine (not restricted to TCM), 5.9% used acupuncture, and 2.5% used TCM (excluding acupuncture) during the past 12 months.[2] Extrapolating these proportions (acupuncture + TCM) to the entire Swiss adult population in 2017,[11] a total of 585,000 Swiss residents underwent at least one TCM-related therapy in the past year by one of the approximately 1000 TCM registered practitioners[12] and 648 registered physicians specialized in TCM in Switzerland.[13] Assuming a regular tariff for a TCM therapy session of 140 CHF, a minimum of 81.9 and 983 Mio. Swiss Francs were spent in 2017 when assuming 1 or 12 therapy session per person, respectively.

Despite the lack of real-world data on TCM usage in Switzerland and most European countries, studies on CM can still provide important complementary information. In Europe, CM was reported to be used among individuals with skin problems (38.1%), back or neck pain (38.0%), allergies (36.7%), stomach or digestive system-related problems (35.7%), upper extremity pain (34.7%), and severe headaches (34.1%). With regard to other health conditions, including cancer and depression, the usage was around 30%.[14] These findings suggest that CM is primarily used to address pain-related issues, which is in accordance with the TCM research activities in Switzerland.

Whether the absolute number of published articles on TCM (N = 223) matches the wide use of TCM in Switzerland remains subjective. When comparing the unfiltered Pubmed literature results using the TCM search term described in the methods (N = 498 articles) with the corresponding search term for Oncology (N = 43,962), Cardiology (N = 14,324) or Pulmonology (N = 6,883), then TCM research seems to be underrepresented. Although this comparison remains superficial, it reveals several hurdles researchers face when conducting TCM research in Switzerland. First – and in contrast to China – only a very few Swiss hospitals systematically apply TCM to their patients; therefore, TCM studies requiring higher numbers of patients have to be conducted in multiple practices or have to be implemented with high costs and hurdles in the hospitals. Second, funding of medical research is often directed to Western Medicine and less to CM or TCM, although exceptions exist, such as the Swiss National Sciences Foundation. These restrictions in patient recruitment and funding opportunities result in single, high-quality, expensive randomized clinical trials in TCM. While it is beneficial to have a few randomized clinical trials instead of many low-quality studies, these single randomized clinical trials are insufficient for a comprehensive and in-depth understanding of TCM usage and its potential benefits for patients in Switzerland and abroad. Moreover, more original, pragmatic, real-world studies, representing the real-world use of TCM across various health conditions, including studies in children, are warranted. In comparison to Europe, where CM therapies are frequently and increasingly used in children,[15] Switzerland could benefit from exploring the feasibility and evidence of using TCM treatments in paediatric populations. Moreover, acupuncture is one of the most commonly researched treatment modalities in paediatric CM therapies.[16–18] Switzerland could further explore the feasibility and evidence of using TCM treatment in children.

This systematic literature review aimed to understand and describe the progression of TCM research activities in relation to the TCM usage in Switzerland, independent of the research activities of other countries in the field of TCM. It is important to note, that findings from TCM studies outside of Switzerland may be applicable to Switzerland, however, in comparison to Western Medicine, TCM’s strength lays in the individual treatment of a patient. Therefore, the transferability and generalizability of findings of TCM studies needs to be better understood. Due to the inclusion of various types of research articles and study designs, this systematic literature review does not include a risk of bias assessment.

## 5. Conclusion

This systematic literature review revealed a gradual increase in TCM research activities in Switzerland, especially since TCM was included in the health insurance reimbursement lists in 2009. However, the absolute number of published articles remains low. Further improvement in TCM knowledge might be achieved by conducting more original studies; elaborating more interdisciplinary and multicentre collaborations; extending TCM research to health conditions treated in TCM practices; investigating other TCM therapies than acupuncture; or focusing on preventive effects of TCM. Obtained knowledge from future original studies conducted in Switzerland will directly and unbiasedly contribute to improved patient care and our understanding of TCM on how to optimally implement and use TCM in the Swiss Western Medicine health system.

## Data Availability

The data underlying the results presented in the study are available from the corresponding author.

## Author contributions

BC, XL, AS and MF contributed to the conception and design of the study. BC, KL, CZ, DJ, SS, HT, WT extracted the data from the articles. BC performed the statistical analysis and wrote the manuscript. All authors critically reviewed and approved the submitted manuscript version.

## References

1. Klein SD, Torchetti L, Frei-Erb M, Wolf U. Usage of Complementary Medicine in Switzerland: Results of the Swiss Health Survey 2012 and Development Since 2007. PLoS One. 2015;10(10):e0141985. Epub 20151029. doi: 10.1371/journal.pone.0141985. PubMed PMID: 26513370; PubMed Central PMCID: PMCPMC4626041.

2. Meier-Girard D, Lüthi E, Rodondi PY, Wolf U. Prevalence, specific and non-specific determinants of complementary medicine use in Switzerland: Data from the 2017 Swiss Health Survey. PLoS One. 2022;17(9):e0274334. Epub 20220914. doi: 10.1371/journal.pone.0274334. PubMed PMID: 36103571; PubMed Central PMCID: PMCPMC9473626.

3. Médecine complémentaire : nouvelles règles de remboursement: Le conseil fédéral admin.ch; 2017 [2024/2/15]. Available from: https://www.admin.ch/gov/fr/accueil/documentation/communiques.msg-id-67050.html.

4. Gang WJ, Xiu WC, Shi LJ, Zhou Q, Jiao RM, Yang JW, et al. Factors Associated with the Magnitude Of acUpuncture treatment effectS (FAMOUS): a meta-epidemiological study of acupuncture randomised controlled trials. BMJ Open. 2022;12(8):e060237. Epub 20220829. doi: 10.1136/bmjopen-2021-060237. PubMed PMID: 36038176; PubMed Central PMCID: PMCPMC9438103.

5. Eisenhardt S, Fleckenstein J. Traditional Chinese medicine valuably augments therapeutic options in the treatment of climacteric syndrome. Arch Gynecol Obstet. 2016;294(1):193–200. Epub 20160404. doi: 10.1007/s00404-016-4078-x. PubMed PMID: 27040419.

6. Blechschmidt T, Krumsiek M, Todorova MG. Acupuncture benefits for Flammer syndrome in individuals with inherited diseases of the retina. Epma j. 2017;8(2):177–85. Epub 20170531. doi: 10.1007/s13167-017-0096-4. PubMed PMID: 28725294; PubMed Central PMCID: PMCPMC5486528.

7. Stöckigt DMB, Kirschbaum B, Carstensen DMM, Witt D, Brinkhaus DMB. Prophylactic Acupuncture Treatment During Chemotherapy in Patients With Breast Cancer: Results of a Qualitative Study Nested in a Randomized Pragmatic Trial. Integr Cancer Ther. 2021;20:15347354211058207. doi: 10.1177/15347354211058207. PubMed PMID: 34814766; PubMed Central PMCID: PMCPMC8646188.

8. Zhu W, Zhang Y, Huang Y, Lu L. Chinese Herbal Medicine for the Treatment of Drug Addiction. Int Rev Neurobiol. 2017;135:279–95. Epub 20170325. doi: 10.1016/bs.irn.2017.02.013. PubMed PMID: 28807162.

9. Zhang XW, Hou WB, Pu FL, Wang XF, Wang YR, Yang M, et al. Acupuncture for cancer-related conditions: An overview of systematic reviews. Phytomedicine. 2022;106:154430. Epub 20220905. doi: 10.1016/j.phymed.2022.154430. PubMed PMID: 36099656.

10. Page MJ, McKenzie JE, Bossuyt PM, Boutron I, Hoffmann TC, Mulrow CD, et al. The PRISMA 2020 statement: an updated guideline for reporting systematic reviews. Bmj. 2021;372:n71. Epub 2021/03/31. doi: 10.1136/bmj.n71. PubMed PMID: 33782057; PubMed Central PMCID: PMCPMC8005924 http://www.icmje.org/conflicts-of-interest/ and declare: EL is head of research for the BMJ; MJP is an editorial board member for PLOS Medicine; ACT is an associate editor and MJP, TL, EMW, and DM are editorial board members for the Journal of Clinical Epidemiology; DM and LAS were editors in chief, LS, JMT, and ACT are associate editors, and JG is an editorial board member for Systematic Reviews. None of these authors were involved in the peer review process or decision to publish. TCH has received personal fees from Elsevier outside the submitted work. EMW has received personal fees from the American Journal for Public Health, for which he is the editor for systematic reviews. VW is editor in chief of the Campbell Collaboration, which produces systematic reviews, and co-convenor of the Campbell and Cochrane equity methods group. DM is chair of the EQUATOR Network, IB is adjunct director of the French EQUATOR Centre and TCH is co-director of the Australasian EQUATOR Centre, which advocates for the use of reporting guidelines to improve the quality of reporting in research articles. JMT received salary from Evidence Partners, creator of DistillerSR software for systematic reviews; Evidence Partners was not involved in the design or outcomes of the statement, and the views expressed solely represent those of the author.

11. Website of the Federal Office for Statistics - Permanent resident population by age and dependency ratio [05.07.2024]. Available from: https://www.bfs.admin.ch/bfs/en/home/statistics/population/effectif-change/age.html.

12. Website of the Swiss TCM therapist Society [05.07.2024]. Available from: https://www.tcm-therapeuten.ch/dir-search/.

13. Website of certified Swiss FMH Doctors [updated 05.07.2024]. Available from: https://www.doctorfmh.ch/en/.

14. Kemppainen LM, Kemppainen TT, Reippainen JA, Salmenniemi ST, Vuolanto PH. Use of complementary and alternative medicine in Europe: Health-related and sociodemographic determinants. Scand J Public Health. 2018;46(4):448–55. Epub 20171004. doi: 10.1177/1403494817733869. PubMed PMID: 28975853; PubMed Central PMCID: PMCPMC5989251.

15. Zuzak TJ, Boňková J, Careddu D, Garami M, Hadjipanayis A, Jazbec J, et al. Use of complementary and alternative medicine by children in Europe: published data and expert perspectives. Complement Ther Med. 2013;21 Suppl 1:S34–47. Epub 20120210. doi: 10.1016/j.ctim.2012.01.001. PubMed PMID: 23578916.

16. Längler A, Zuzak TJ. Complementary and alternative medicine in paediatrics in daily practice-- a European perspective. Complement Ther Med. 2013;21 Suppl 1:S26–33. Epub 20120302. doi: 10.1016/j.ctim.2012.01.005. PubMed PMID: 23578914.

17. Huber BM, von Schoen-Angerer T, Hasselmann O, Wildhaber J, Wolf U. Swiss paediatrician survey on complementary medicine. Swiss Med Wkly. 2019;149:w20091. Epub 20190616. doi: 10.4414/smw.2019.20091. PubMed PMID: 31203577.

18. Piñeiro Pérez R, Núñez Cuadros E, Cabrera García L, Díez López I, Escrig Fernández R, Gil Lemus M, et al. Results of a national survey on knowledge and use of complementary and alternative medicine by paediatricians. An Pediatr (Engl Ed). 2022;96(1):25–34. Epub 20211211. doi: 10.1016/j.anpede.2020.09.012. PubMed PMID: 34906426.

